# Changes in intake of dairy product subgroups and risk of type 2 diabetes: modelling specified food substitutions in the Danish Diet, Cancer and Health cohort

**DOI:** 10.1101/2020.05.28.20116459

**Authors:** Daniel B. Ibsen, Kim Overvad, Anne Sofie D. Laursen, Jytte Halkjær, Anne Tjønneland, Tuomas O. Kilpeläinen, Erik T. Parner, Marianne U. Jakobsen

**Affiliations:** Department of Public Health, Aarhus University, Aarhus, Denmark; Danish Cancer Society Research Center, Copenhagen, Denmark; Department of Public Health, University of Copenhagen, Copenhagen, Denmark; Novo Nordisk Foundation Center for Basic Metabolic Research, Faculty of Health and Medical Sciences, University of Copenhagen, Copenhagen, Denmark; National Food Institute, Division for Diet, Disease Prevention and Toxicology, Technical University of Denmark, Kgs. Lyngby, Denmark

**Keywords:** diet change, substitution models, follow-up studies, dairy, diabetes

## Abstract

We investigated the association between an increased intake of one dairy product subgroup at the expense of another within a 5-year period and the subsequent 10-year risk of developing type 2 diabetes. Effect modification by the initial level of the dairy product intake was also examined. The cohort included 39,438 men and women with two measurements of diet assessed using food frequency questionnaires (FFQ) administered in 1993-1997 and 1999-2003. Dairy products were skimmed milk, semi-skimmed milk, whole-fat milk, buttermilk, low-fat yogurt, whole-fat yogurt, cheese and butter. Type 2 diabetes cases were ascertained from the Danish National Diabetes Register. The pseudo-observation method was used to calculate risk differences (RD) with 95% confidence intervals (CI). Among participants aged 56-59 years at hand-in of the follow-up FFQ, increased intake of whole-fat yogurt in place of skimmed, semi-skimmed or whole-fat milk was associated with a reduced risk (RD% [95% CI]: −0.8% [−1.3, −0.2]; −0.6% [−1,1, −0.1]; −0.7 [−1.2, −0.1]; per 50g/d, respectively). Among participants aged 60-64 and 65-72, substitution of skimmed milk for semi-skimmed milk was associated with an increased risk of type 2 diabetes (0.5% [0.2, 0.7]; 0.4% [0.1, 0.7]; per 50g/d, respectively). No effect modification by the initial intake of milk products was observed. Our results suggest that substitution of whole-fat yogurt for milk among those aged 56-59 decreases the risk of type 2 diabetes and substitution of skimmed milk for semi-skimmed milk was associated with an increased risk of type 2 diabetes among those aged 60-64 and 65-72, regardless of the initial intake level.

## Introduction

The incidence of type 2 diabetes is increasing rapidly worldwide [1] and the prevention of type 2 diabetes is a top priority in public health [2]. Diet, together with other lifestyle factors, plays a key role in prevention of type 2 diabetes. Indeed, risk of type 2 diabetes has been found to remain lower at least 10 years after participation in lifestyle interventions that included dietary modifications lasting two to six years [3].

Current dietary recommendations to the general population [4,5] and for prevention of type 2 diabetes [6,7] recommend low-fat dairy products instead of whole-fat dairy products [8]. However, meta-analyses of randomized interventions have found no clear effect of increasing the total intake of dairy products on glucose metabolism or insulin sensitivity and no effect of increasing the intake of low-fat or high-fat dairy products on cardio-metabolic risk markers, expect for a slightly increased body weight [9–11]. This lack of an effect may be due to small sample sizes and short durations of the included trials and to differences in types of dairy products provided and control groups used, either consuming their habitual diet, a low-dairy or no-dairy diet, between the included trials. Only few cohort studies have investigated the association between changes in intake of dairy products on the risk of developing type 2 diabetes [12]. Of the few cohort studies that investigated changes in intake of dairy products, higher consumption of yogurt, not milk products, was associated with a lower risk of type 2 diabetes [13]. To be sure that assessed associations are independent of changes in energy intake, it is important to adjust for energy intake in the statistical model. Consequently, a food substitution is introduced where the model now reflects the influence of increasing the intake of, for example, yogurt products, as in the example, at the expense of a mix of other foods on the risk of type 2 diabetes; mimicking an isocaloric intervention study. As all foods are either harmful, neutral or beneficial in the prevention of type 2 diabetes, the association between intake of a specific dairy product subgroup and the risk of type 2 diabetes depend on the substituted food. Unfortunately, this has rarely been specified, resulting in an unclear interpretation of the results.

Another relevant question when investigating dietary changes is whether the initial intake modifies the association between a dietary change and development of disease. In the context of changes in dairy product subgroups, the association between substitution of, for instance, skimmed milk for whole-fat milk and the risk of type 2 diabetes may be different, depending on whether the initial intake of whole-fat milk was high or low.

Specifying food substitutions when studying associations between dietary changes and long-term disease risk may add important new knowledge on the public health impact of changing diet. Thus, we investigated the association between an increased intake of one dairy product subgroup at the expense of another within a 5-year period, and the subsequent 10-year risk of developing type 2 diabetes in middle-aged men and women. In addition, we examined whether the association was modified by the initial intake of these products.

## Materials and methods

### Study population

The Diet, Cancer and Health cohort is a Danish cohort that recruited participants from 1993-1997. Middle-aged men and women from the Aarhus and Copenhagen areas were invited to participate in the study. At inclusion in 1993-1997 participants answered a validated food frequency questionnaire (FFQ) mailed to the participants [14,15]. Furthermore, the participants underwent a physical examination and answered a lifestyle questionnaire at one of two study centres [16]. In 1999-2003, participants from the original cohort answered mailed follow-up questionnaires on diet and lifestyle similar to the questionnaires provided in 1993-1997.

For the purposes of the present study, participants with a diagnosis of diabetes, myocardial infarction, stroke or cancer before the time of the hand-in of questionnaires in 1999-2003 were excluded. Furthermore, participants for whom we did not have information on exposures or covariates were also excluded.

### Dairy product subgroups

The initial intake of dairy product subgroups (skimmed milk 0.1% fat, semi-skimmed milk 1.5%, whole-fat milk 3.5%, buttermilk 0.5%, low-fat yogurt < 1.5%, whole-fat yogurt 3.5%, cheese and butter) was assessed in 1993-1997 using a validated FFQ designed for the Diet, Cancer and Health cohort [14,15]. The FFQ inquired participants about their average intake of the dairy product subgroups during the previous 12 months according to 12 possible categories ranging from ‘never’ to ‘eight times or more per day’. Low-fat yogurt and whole-fat yogurt included plain yogurt types and yogurt with fruit. Participants handed in the FFQ at one of the two study centres and it was checked by trained personnel that immediately addressed inconsistencies of the questionnaires with the participants, if necessary. In 1999-2003, a FFQ similar to the 1993-1997 FFQ was mailed to the participants and the returned FFQs were checked by trained personnel. Missing values were scored in terms of the type and relevance of the question (0, 0.5, 1 or 10 points). Participants with > 9 points were contacted by phone. If participants had > 55 points, the FFQ was mailed back to the participants. The remaining missing values were coded as the lowest intake level “never/seldom”. Harmonization of the dietary data collected in 1993-1997 and 1999-2003 is described in the Supplemental text. Food and nutrient intakes in g/day were calculated using FoodCalc [17] and Danish food composition tables [18] version 1996. Changes in intake of dairy product subgroups were defined as the difference in reported intake in 1993-1997 (initial intake) and reported intake in 1999-2003, and expressed in units of one serving (50 g/day for skimmed milk, semi-skimmed milk, whole-fat milk, buttermilk, low-fat and whole-fat yogurt, 20g/day for cheese and 6 g/day for butter).

### Type 2 diabetes ascertainment

Type 2 diabetes status of the participants was ascertained from the Danish National Diabetes Register [19]. This register combines information from several Danish registers and covers the entire Danish population. The date of diagnosis was the earliest date registered with (a) a diagnosis of diabetes in the Danish National Patient Register; (b) chiropody as a diabetes patient in the Danish National Health Service Register; (c) five blood glucose measurements in a 1-year period or two blood glucose measurements per year in five consecutive years in the Danish National Health Service Register; or (d) purchase of oral anti-diabetes drugs in the Danish National Prescription Registry (excluding females prescribed metformin alone). The algorithm has been validated with a positive predictive value of 89% as compared to medical records from the general practitioner [19]. The register does not distinguish between type 1 and type 2 diabetes. However, as all participants in the Diet, Cancer and Health cohort were middle-aged, the vast majority of incident cases is expected to be type 2 diabetic cases. The participants were followed up from the hand-in of questionnaires in 1999-2003 until the date of the diagnosis of type 2 diabetes, death, emigration or the date at which participants had been followed for 10 years after hand-in of questionnaires in 1999-2003, whichever came first. It is not known when the participants changed their dietary intake during the 5 years between the two FFQs, but 10 years of subsequent follow-up was considered reasonable based on evidence from clinical trials showing a lower risk of type 2 diabetes for up to 10 years after initiation of lifestyle interventions lasting two to six years [3].

### Assessment of covariates

Information on covariates was obtained from the lifestyle questionnaires and FFQs administered in 1993-1997 and 1999-2003. The analyses were adjusted for known risk factors of type 2 diabetes selected a priori based on a review of the literature. From the 1993-1997 questionnaires, information on education, history of hypertension, history of hypercholesterolemia, total energy intake and food groups associated with type 2 diabetes was obtained. From the questionnaires in 1999-2003, information on alcohol intake, smoking status, physical activity, body weight and waist circumference and updated information on history of hypertension, history of hypercholesterolemia, total energy intake and food groups was obtained. In addition, participants self-reported their family history of diabetes. Height was measured at the physical examination in 1993-1997.

### Statistical analysis

We calculated medians and 80% central ranges for continuous variables, and proportions for categorical variables among the cohort and cases.

We used the pseudo-observation method [20,21] with age as the underlying time-scale to calculate risk differences (RDs) and their corresponding 95% confidence intervals (CIs). A central assumption of the pseudo-observation method is that there must be marginally independent entry and censoring. We investigated the validity of this assumption using a previously described methodology stratifying the cohort based on age and date of inclusion, which was the date at hand-in of the questionnaires in 1999-2003 [22]. Because age was the underlying-time scale, a specific age was set as the end of the follow-up period as required for the generation of the pseudo-values. We used the median age at which participants were followed-up for 10 years after hand-in of questionnaires in 1999-2003 as the end of the risk period. Risk periods were 56-67 years, 60-71 years and 65-76 years, in stratum 1, 2 and 3, respectively. Death during follow-up was handled as a competing risk. The substitution models included all dairy product subgroups as change variables and for all dairy product subgroups assessed in 1993-1997. The difference between two change-variables’ beta-coefficients (the beta-coefficient for the product increased minus the beta-coefficient for the product decreased) and their variances and covariance were used to estimate the RDs and corresponding 95% CIs for a given substitution. Hence, the association of an increased intake of a specific dairy product subgroup and a concomitant decreased intake of another specified dairy product subgroup (i.e. a substitution) in the 5-year period from 1993-1997 to 1999-2003, with the subsequent 10-year risk of developing type 2 diabetes, was estimated. In multivariable analyses we included total energy intake (1993-1997 and 1999-2003, kJ/day; continuous) and sex (man, woman) in Model 1a. Total energy intake at both time-points was included to assess the association between substitutions independent of total energy intake at both time-points and to reduce measurement error of reported food intake [23]. Consequently, an unspecified residual substitution was introduced, which is equal to the difference in energy provided by the substituted products. Model 1b was further adjusted for the duration of education (≤7, 8-10, > 10 years), assessed in 1993-1997, and alcohol intake (g/day; continuous, as a restricted cubic spline with three knots), smoking status (never, former, current < 15 g tobacco/day, current 15-25 g tobacco/day, current > 25 g tobacco/day), physical activity (participating in leisure-time sport activities or not), and family history of diabetes (yes, no, do not know) assessed in 1999-2003. Model 2 was further adjusted for other foods associated with type 2 diabetes: fruit, vegetables, red and processed meat, sugar-sweetened beverages, whole grains, potato chips and coffee (all in g/day; continuous) assessed in 1999-2003. Lastly, body mass index (BMI) (kg/m^2^; continuous), waist circumference (cm; continuous), history of hypertension (yes, no, do not know) and history of hypercholesterolemia (yes, no, do not know), assessed in 1999-2003, were added in a separate model (Model 3), as these covariates may be considered as mediators rather than confounders. Linearity for the dairy product variables was investigated using restricted cubic spline of each dairy product subgroup. We constructed a correlation matrix of dairy product intake, assessed in 1993-1997, and changes in dairy product intake in order to assess potential multicollinearity (i.e. one variable almost completely explains another variable) as changes in dairy product variables may be highly correlated with the initial intake of dairy product subgroups.

We investigated potential effect modification by the initial intake level of the replaced dairy product group assessed in 1993-1997 in subgroup analyses. For instance, the association between substitution of skimmed milk for whole-fat milk and risk of type 2 diabetes in subgroup analyses by initial low or high whole-fat milk intake assessed in 1993-1997 (cut at the 50th percentile). We also conducted sex-stratified analyses.

As a sensitivity analysis, we excluded participants who developed hypertension and/or hypercholesterolemia between 1993-1997 and 1999-2003. This was done because these participants are likely to have changed their dietary intake as a result of diagnosis and treatment. Furthermore, non-participation at follow-up may result in selection bias. Thus, in another sensitivity analysis, we adjusted for potential selection bias by using inverse probability weighting [24]. The probability weights were based on sex, age, education, alcohol, smoking, physical activity, BMI, waist circumference, history of hypertension, history of hypercholesterolemia, history of diabetes and alternate healthy eating index-2010 score [25] assessed in 1993-1997. Also, we compared characteristics assessed with the 1993-1997 questionnaires between those who participated at both time-points and those who only participated in 1993-1997. Changing two dietary components may be followed by other dietary changes as well. We constructed spider plots to visually assess differences in changes in intake of different food groups between participants that substituted one for another dairy product subgroup, compared with participants who maintained a stable intake of both dairy product subgroups. Substitutions were defined as increased intake of one dairy product subgroup by ≥1 serving/day and decreased intake of another dairy product subgroup by ≥1 serving/day. To investigate the time-scale sensitivity of our results, we compared risk ratios from the analyses with age as the underlying time-scale with risk ratios from analyses using time in the study as the underlying time-scale. We compared risk ratios and not RDs as risk ratios are generally less sensitive across studies than risk differences. Hence, differences between time-scales using risk ratios would be of a greater concern than dissimilarities using risk differences. Lastly, we excluded 7179 participants with > 9 missing values in their 1999-2003 FFQ coded as “no intake or less than once per month” in the main analysis.

All analyses were performed in Stata version 15.1 (StataCorp, College Station, Texas, USA). Forest plots were, however, coded in R version 3.5.0, R Core Team, Vienna, Austria, using the ggepi package [26].

## Results

In total, two measures of dietary intake were available for 45,247 participants (79% of the initial cohort) (Figure 1). We excluded individuals who had a diagnosis of diabetes (n = 1971), myocardial infarction or stroke (n = 1518), or cancer (n = 1621) before hand-in of the questionnaires in 1999-2003, and participants for whom there were missing values in exposures or covariates (n = 699), leaving 39,438 eligible participants.

**Figure 1.**
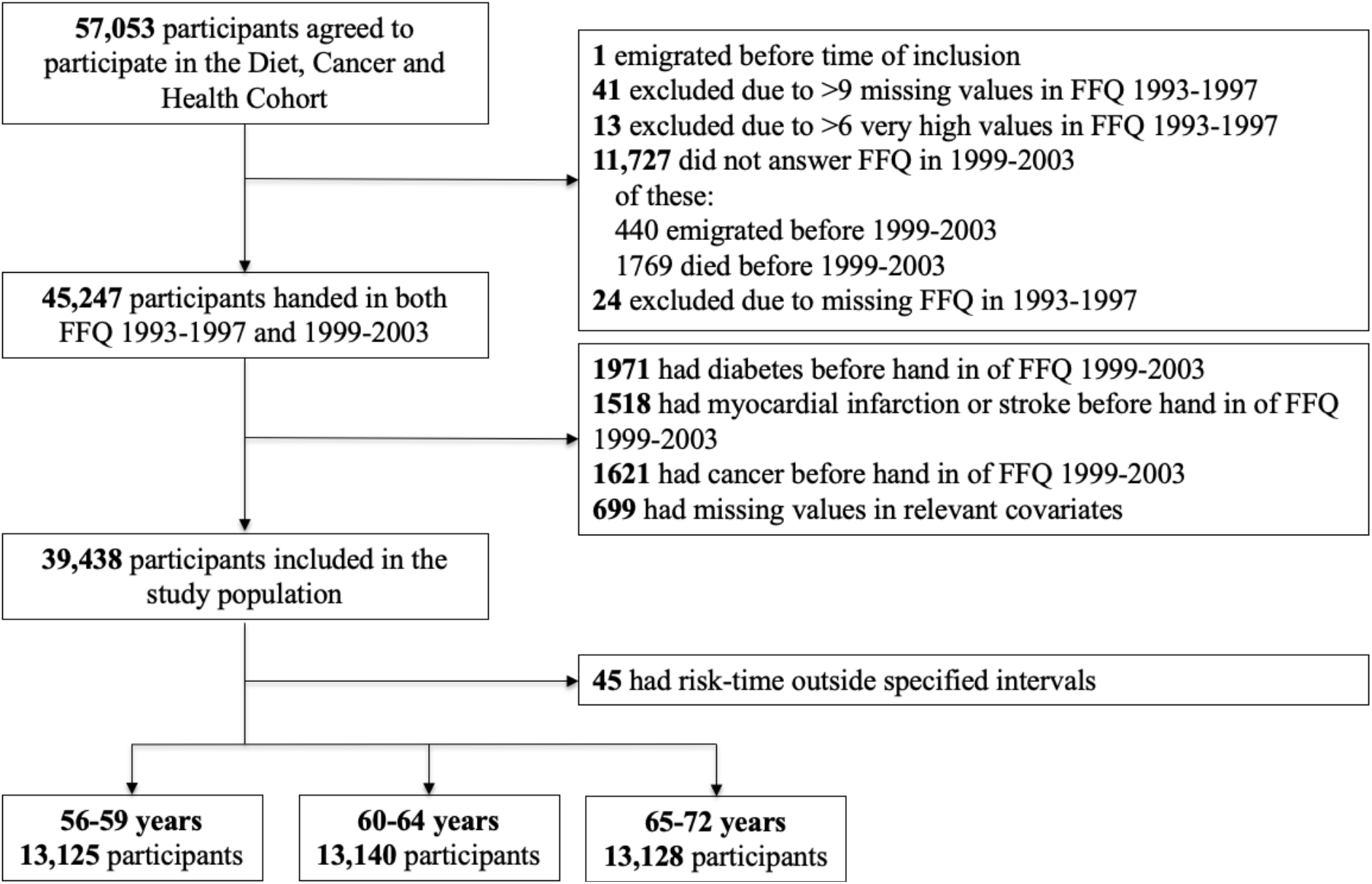
Flowchart of the study sample in the Diet, Cancer and Health cohort. FFQ: food frequency questionnaire.

The assumption of independent entry within the entire cohort did not hold (hazard ratio [95% CI] for type 2 diabetes comparing first and third tertile of age at inclusion: 1.26 [1.11, 1.42]). Based on this result and a previous study [27], we analysed substitutions within three strata based on age at hand-in of the questionnaires in 1999-2003 and three strata based on calendar date at hand-in of the questionnaires in 1999-2003 (in total 9 strata). Within each stratum, no violation of the assumptions was observed. Thus, the pseudo-values were generated in each of these strata. Because the pseudo-values can only be combined if they span over the same time-period within the used time-scale, the data were analysed in three separate age-strata. Within each age-stratum, calendar date-strata were combined after generation of pseudo-values with similar start and end ages for cumulative risks within each age-stratum. However, as a consequence, we excluded 45 individuals with risk-time outside these specified intervals. Stratum 1, aged 56-59 years, included 13,125 participants of which 1043 developed diabetes, stratum 2, 60-64 years, included 13,140 participants and 1188 cases and stratum 3, 65-72 years, included 13,128 participants and 1281 cases. The end of each follow-up period was set at the ages 67, 71 and 76 years with a median follow-up time of 9.5, 9.1 and 8.3 years for strata 1, 2 and 3, respectively.

Those who developed type 2 diabetes were more likely to be less educated, be smokers, not participate in leisure-time sports activities, have a higher BMI and waist circumference and have a history of hypertension or hypercholesterolemia and a family history of diabetes at hand-in of the follow-up questionnaires (Table 1). There were no substantial differences in the median changes in intake of dairy product subgroups between the cohort and those who developed type 2 diabetes. However, the ranges of changes in intake (p10-p90) were different between the two (Table 1).

**Table 1.**
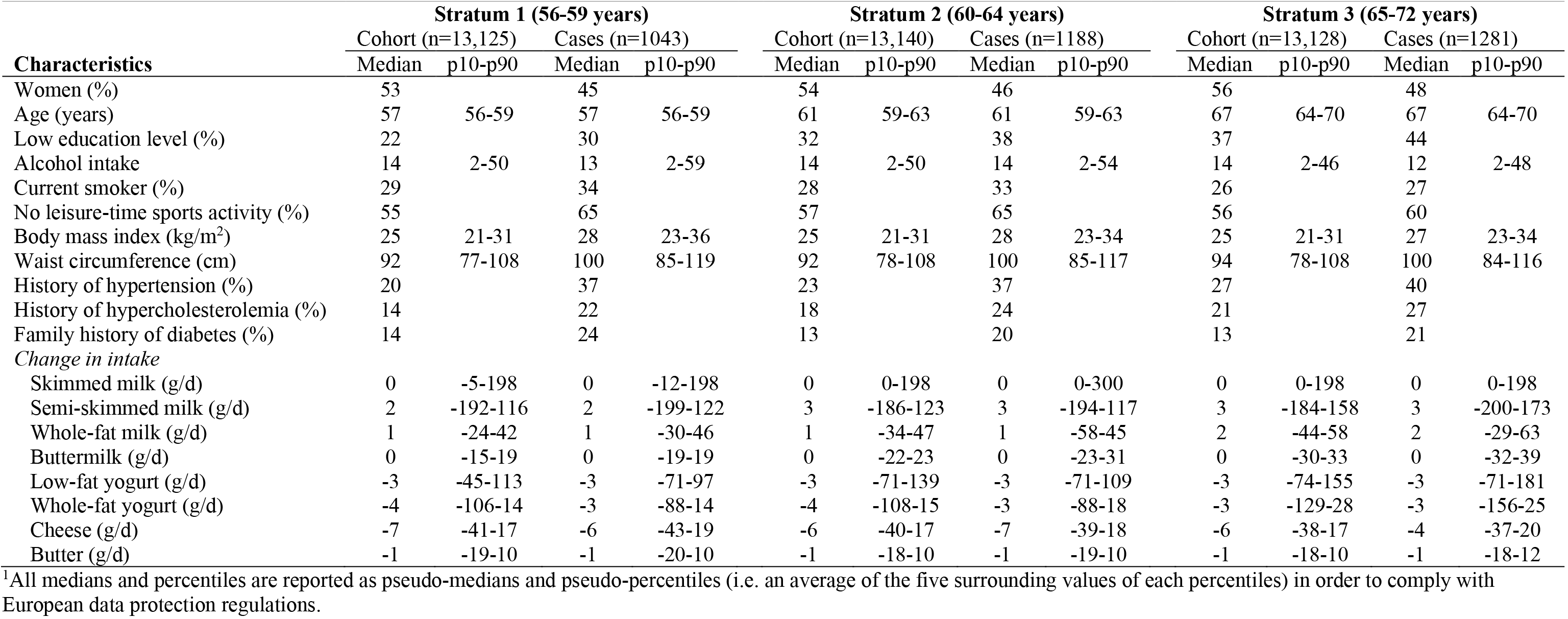
Characteristics at hand-in of follow-up questionnaires (1999-2003) and changes in intake of dairy product subgroups of participants in the Diet, Cancer and Health cohort in three strata divided into tertiles of age^1^.

| Characteristics | Stratum 1 (56-59 years) |  |  |  | Stratum 2 (60-64 years) |  |  |  | Stratum 3 (65-72 years) |  |  |  |
| --- | --- | --- | --- | --- | --- | --- | --- | --- | --- | --- | --- | --- |
|  | Cohort (n=13,125) |  | Cases (n=1043) |  | Cohort (n=13,140) |  | Cases (n=1188) |  | Cohort (n=13,128) |  | Cases (n=1281) |  |
|  | Median | p10-p90 | Median | p10-p90 | Median | p10-p90 | Median | p10-p90 | Median | p10-p90 | Median | p10-p90 |
| Women (%) | 53 |  | 45 |  | 54 |  | 46 |  | 56 |  | 48 |  |
| Age (years) | 57 | 56-59 | 57 | 56-59 | 61 | 59-63 | 61 | 59-63 | 67 | 64-70 | 67 | 64-70 |
| Low education level (%) | 22 |  | 30 |  | 32 |  | 38 |  | 37 |  | 44 |  |
| Alcohol intake | 14 | 2-50 | 13 | 2-59 | 14 | 2-50 | 14 | 2-54 | 14 | 2-46 | 12 | 2-48 |
| Current smoker (%) | 29 |  | 34 |  | 28 |  | 33 |  | 26 |  | 27 |  |
| No leisure-time sports activity (%) | 55 |  | 65 |  | 57 |  | 65 |  | 56 |  | 60 |  |
| Body mass index (kg/m<sup>2</sup>) | 25 | 21-31 | 28 | 23-36 | 25 | 21-31 | 28 | 23-34 | 25 | 21-31 | 27 | 23-34 |
| Waist circumference (cm) | 92 | 77-108 | 100 | 85-119 | 92 | 78-108 | 100 | 85-117 | 94 | 78-108 | 100 | 84-116 |
| History of hypertension (%) | 20 |  | 37 |  | 23 |  | 37 |  | 27 |  | 40 |  |
| History of hypercholesterolemia (%) | 14 |  | 22 |  | 18 |  | 24 |  | 21 |  | 27 |  |
| Family history of diabetes (%) | 14 |  | 24 |  | 13 |  | 20 |  | 13 |  | 21 |  |
| *Change in intake* |  |  |  |  |  |  |  |  |  |  |  |  |
| Skimmed milk (g/d) | 0 | -5-198 | 0 | -12-198 | 0 | 0-198 | 0 | 0-300 | 0 | 0-198 | 0 | 0-198 |
| Semi-skimmed milk (g/d) | 2 | -192-116 | 2 | -199-122 | 3 | -186-123 | 3 | -194-117 | 3 | -184-158 | 3 | -200-173 |
| Whole-fat milk (g/d) | 1 | -24-42 | 1 | -30-46 | 1 | -34-47 | 1 | -58-45 | 2 | -44-58 | 2 | -29-63 |
| Buttermilk (g/d) | 0 | -15-19 | 0 | -19-19 | 0 | -22-23 | 0 | -23-31 | 0 | -30-33 | 0 | -32-39 |
| Low-fat yogurt (g/d) | -3 | -45-113 | -3 | -71-97 | -3 | -71-139 | -3 | -71-109 | -3 | -74-155 | -3 | -71-181 |
| Whole-fat yogurt (g/d) | -4 | -106-14 | -3 | -88-14 | -4 | -108-15 | -3 | -88-18 | -3 | -129-28 | -3 | -156-25 |
| Cheese (g/d) | -7 | -41-17 | -6 | -43-19 | -6 | -40-17 | -7 | -39-18 | -6 | -38-17 | -4 | -37-20 |
| Butter (g/d) | -1 | -19-10 | -1 | -20-10 | -1 | -18-10 | -1 | -19-10 | -1 | -18-10 | -1 | -18-12 |

<sup>1</sup>All medians and percentiles are reported as pseudo-medians and pseudo-percentiles (i.e. an average of the five surrounding values of each percentiles) in order to comply with European data protection regulations.

For the main substitution analyses we found different patterns across the age-strata (Figure 2 and Supplemental Figure 1). After multivariable adjustment (Figure 2), an increased risk of type 2 diabetes was observed when skimmed milk replaced semi-skimmed milk among those aged 60-64 (RD [95% CI]: 0.5% [0.2, 0.7] per 50 g/d) and 65-72 (RD [95% CI]: 0.4% [0.1, 0.7] per 50 g/d) and when skimmed milk replaced whole-fat milk (RD [95% CI]: 0.4% [0.0, 0.8] in those aged 60-64 and 0.4 [0.0, 0.7] in those aged 65-72). When whole-fat yogurt replaced milk products, a reduced risk of type 2 diabetes was observed among those aged 56-59 (RD [95% CI]: whole-fat yogurt for skimmed milk: −0.8% [−1.3, −0.2]; semi-skimmed milk: −0.6% [−1.1, −0.1]; whole-fat milk: −0.7% [−1.2, −0.1]; per 50 g/d). Replacing skimmed milk, semi-skimmed milk or whole-fat milk with low-fat yogurt was associated with a reduced risk of type 2 diabetes (RD [95% CI]: −0.4% [−0.9, 0.0]; −0.3% [−0.7, 0.2]; −0.3% [−0.8, 0.1]; per 50 g/day, respectively), although not statistically significantly so. In contrast, replacing low-fat yogurt with semi-skimmed or whole-fat milk was associated with an increased risk in those aged 65-72 years (RD [95% CI]: 0.6% [0.1, 1.1]; 0.6% [0.0, 1.1]; per 50 g/day, respectively). The same pattern of associations was found after adjustment for BMI, waist circumference, history of hypertension and hypercholesterolemia (Supplemental Figure 1, Model 3). No deviation from linearity of the dairy product variables and no multicollinearity was detected during the model validation (data not shown).

**Figure 2.**
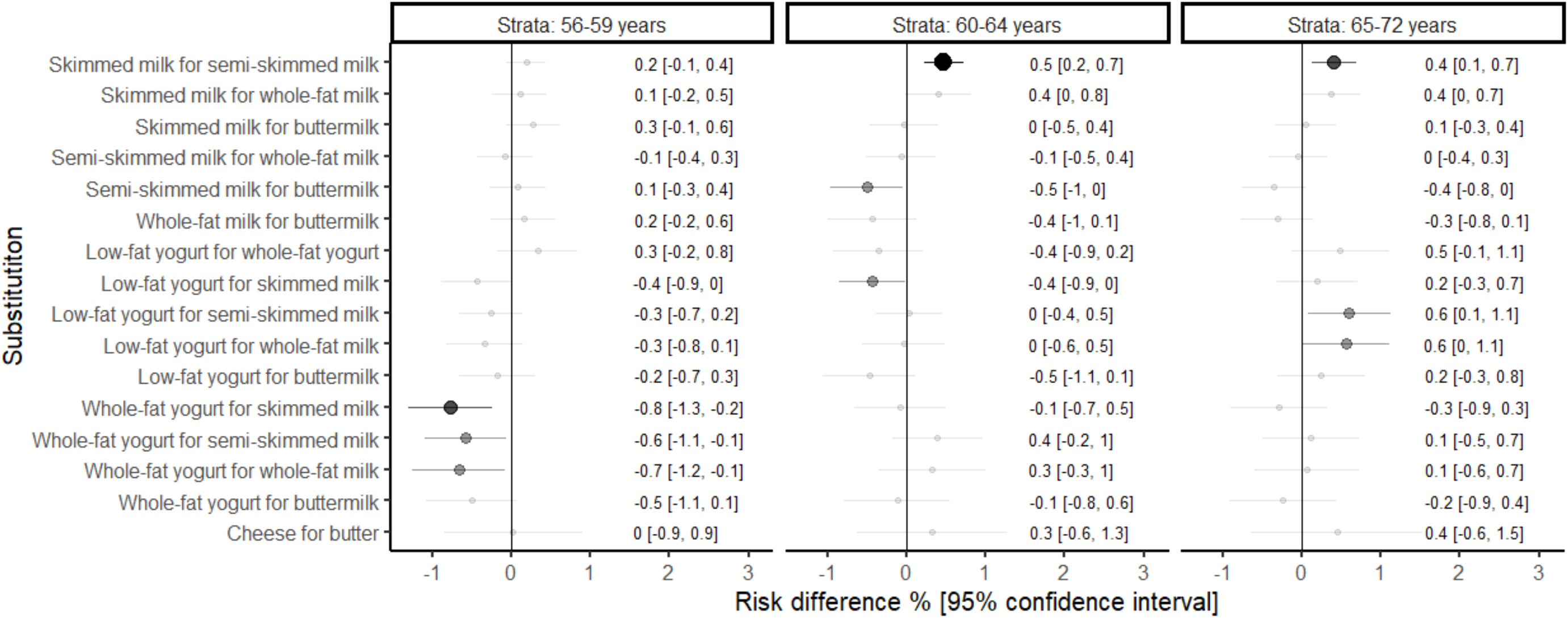
Forest plot of substitutions between dairy product subgroups on 10 years risk of type 2 diabetes in the Diet, Cancer and Health cohort across three strata divided into tertiles of age at hand-in of questionnaires in 1999-2003. 10-year risk periods were 56-67 years, 60-71 years and 65-76 years, in strata 56-59, 60-64 and 65-72 years, respectively. Intakes were modelled per 50 g/day for all dairy products except for cheese (20 g/day) or butter (6 g/day). Adjusted for Model 2 covariates: total energy intake (assessed in 1993-1997 and 1999-2003), sex, duration of education (assessed in 1993-1997) and alcohol intake, smoking status, leisure-time sports activity and family history of diabetes, fruit, vegetables, red and processed meat, sugar-sweetened beverages, whole grains, potato chips and coffee (assessed in 1999-2003). The larger and darker the dots, the lower the p-value for a given association.

Figure 3 shows the associations for substitutions between dairy product subgroups and risk of type 2 diabetes stratified by the initial intake level of the replaced dairy product group. The patterns of associations were largely similar between those with an initial low and high intake of the substituted dairy product subgroup. When the analyses were stratified by sex, no associations between substitutions of whole-fat yogurt for milk and risk of type 2 diabetes were observed among women aged 56-59 whereas substitution of whole-fat yogurt for milk was associated with a reduced risk in men aged 56-59. Substitution of skimmed milk for semi-skimmed milk was associated with an increased risk among women aged 60-64 and 65-72 and only among men aged 60-64 (Supplemental Figure 2).

**Figure 3.**
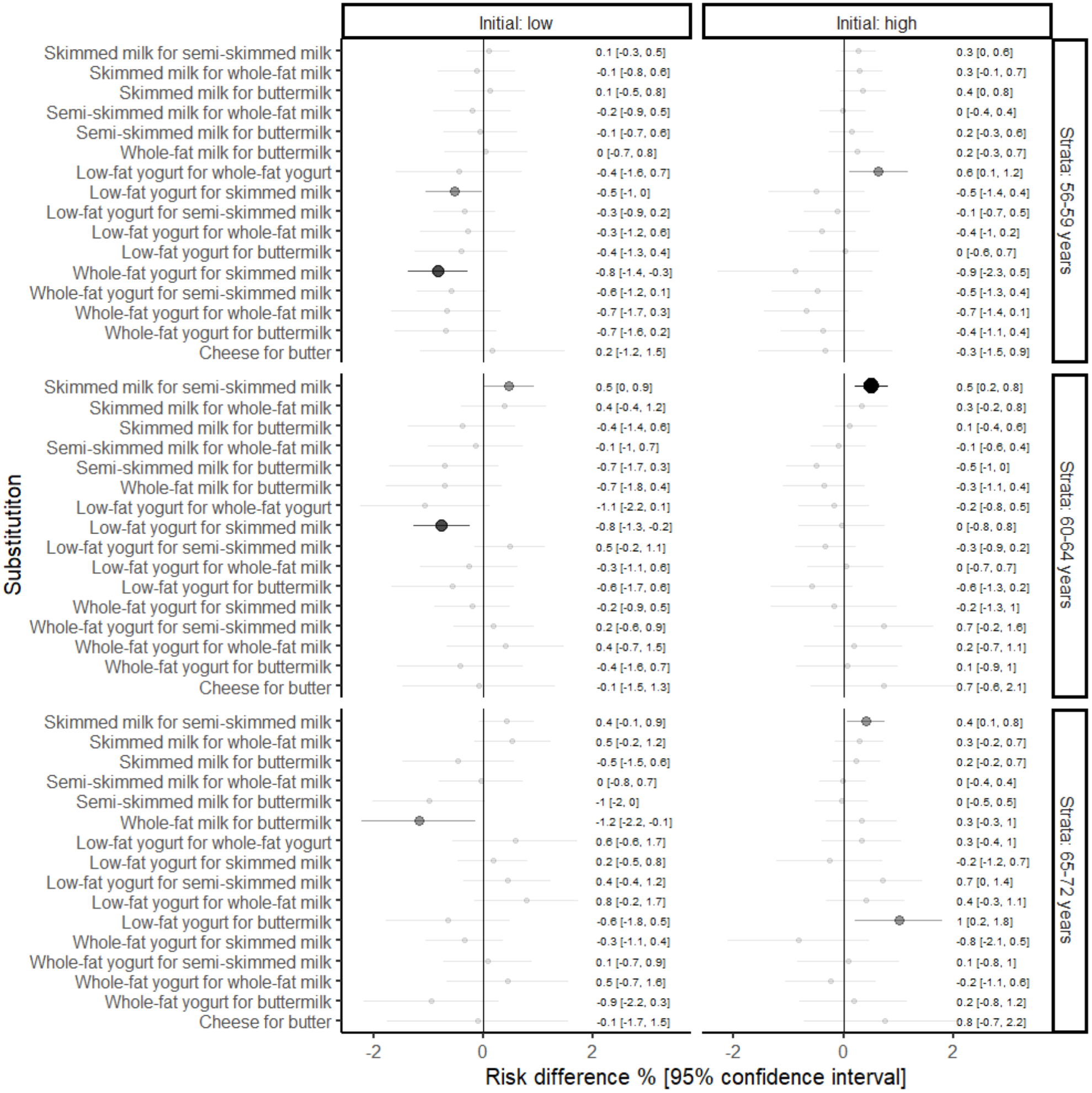
Forest plot of substitutions between dairy product subgroups on risk of type 2 diabetes, followed-up for subsequent 10 years, in the Diet, Cancer and Health cohort across three strata divided into tertiles of age at hand-in of questionnaires in 1999-2003 and low or high initial intake of the substituted dairy product, cut at the 50th percentile. 10-year risk periods were 56-67 years, 60-71 years and 65-76 years, in strata 56-59, 60-64 and 65-72 years, respectively. Intakes were modelled per 50 g/day for all dairy products except for cheese (20 g/day) or butter (6 g/day). Adjusted for Model 2 covariates: total energy intake (assessed in 1993-1997 and 1999-2003), sex, duration of education (assessed in 1993-1997) and alcohol intake, smoking status, leisure-time sports activity and family history of diabetes, fruit, vegetables, red and processed meat, sugar-sweetened beverages, whole grains, potato chips and coffee (assessed in 1999-2003). The larger and darker the dots, the lower the p-value for a given association.

After excluding participants who developed hypertension or hypercholesterolemia between the two dietary assessments, replacement of milk products with yogurt became more strongly associated with a reduced risk of type 2 diabetes (Supplemental Figure 3). Otherwise, similar results as in the main analyses were observed. Supplemental Figure 4 shows the flow of participants that participated both in 1993-1997 and in 1999-2003 and those who only participated in 1993-1997. Non-participants (i.e. those who only participated in 1993-1997) were more likely to be younger, have a lower education, consume less alcohol, be smokers, be less physically active, have a higher BMI and waist circumference and have a lower healthy eating index score in 1993-1997 compared to those who answered both 1993-1997 and 1999-2003 questionnaires (Supplemental Table 1). Adjusting for non-participation using inverse probability weighting provided similar results as the main analyses (Supplemental Figure 5). Supplemental Figure 6 illustrates differences in intake of other foods related to type 2 diabetes comparing participants that have increased their intake of ≥1 serving/day of, for example skimmed milk, and decreased their intake of ≥1 serving/day of, for example semi-skimmed milk (i.e. substituted skimmed milk for semi-skimmed milk), with individuals that did not change their intake of the dairy product subgroups. Subjects that substituted low-fat dairy products, particularly skimmed milk or low-fat yogurt for other dairy products, increased their intake of fruit and vegetables to a greater extent than those with a stable intake. Comparison of different underlying time-scales and exclusion of participants with > 9 missing values in the 1999-2003 FFQ revealed similar patterns of associations compared with the main analyses (Supplemental Figure 7 and 8, respectively).

## Discussion

In this large cohort of middle-aged Danish men and women with repeated measures of diet, we found different patterns of associations across age-strata for specified substitutions of dairy product subgroups. We found that replacing milk products with whole-fat yogurt reduced the risk of type 2 diabetes among those aged 56-59 years. Replacing semi-skimmed milk with skimmed milk was associated with an increased risk of type 2 diabetes among those aged 60-64 and 65-72 years. No clear associations were observed for replacements with buttermilk, cheese or butter.

The primary exposures of this study were changes in intake of dairy product subgroups. To investigate changes we used two measures of diet. The dietary intakes were self-reported and thus measurement error is present. However, the FFQ was validated for nutrient intake and similar FFQs, validated on a food level, tend to show a high correlation with food records for dairy products (r = 0.53-0.90) [28-30]. We thoroughly adjusted for established risk factors of type 2 diabetes, but residual confounding cannot be excluded. Furthermore, after excluding those who developed hypertension or hypercholesterolemia between 1993-1997 and 1999-2003, replacement of milk with yogurt became more strongly associated with a reduced risk of type 2 diabetes. This raise concerns about potential confounding by indication. However, the point in time when the change in diet and development of hypertension or hypercholesterolemia was unknown. To our surprise, we observed an increased risk of type 2 diabetes when replacing semi-skimmed milk with skimmed milk among those aged 60-64 and 65-72 years. This is not consistent with the current evidence, which mostly suggests no association between intake of milk, regardless of fat content, and type 2 diabetes risk [12,31]. Other indicators of deteriorating health status (such as physical disability or chronic inflammation) were not taken into account and thus, we cannot exclude the possibility of confounding by indication here as well. All Danish men and women between ages 50-64 years, without any registered cancer diagnosis, living in the two largest cities in Denmark were invited to participate in the Diet, Cancer and Health cohort. It has previously been shown that those who chose to participate generally had a higher socioeconomic position [16], which limits the generalizability of our results. We observed similar tendencies among participants eligible for this study. Participants had longer duration of education than non-participants. Hence, non-participation at follow-up of about 17% may have introduced selection bias. However, after using inverse probability weights of participation in our analyses, we found similar results compared to the main analyses, suggesting that selection bias is not a major concern.

Only few previous studies have investigated changes in intake of dairy product subgroups on risk of developing type 2 diabetes. Interestingly, we found that replacing milk with yogurt products was associated with a reduced risk of type 2 diabetes in those aged 56-59, but not those aged 60-72 years. As the effect of dietary intake on future risk of developing type 2 diabetes is likely cumulative, changes among older populations may not have the same impact as dietary changes among younger populations. For instance, although intake of fruit and vegetables have been consistently associated with a lower risk of type 2 diabetes [32], these associations were not found in a meta-analysis of cohorts of elderly individuals (>60 years) [33]. Alternatively, higher baseline hazards among the elderly may also explain the findings from this meta-analysis and none of the included studies investigated changes in intake of fruit and vegetables. Current dietary recommendations emphasize substitution of high-fat dairy product with low-fat dairy products, although few cohort studies have investigated the health impact of changes in intake of dairy products. In a cohort of Spanish men and women aged 55-80 years, Diaz-Lopez *et al*. [13], reported that an increased intake of low-fat and whole-fat yogurt from below the median intake to more than or equal to the median intake was associated with a lower risk of type 2 diabetes compared with a stable intake below the median, whereas no similar association was observed for low-fat or whole-fat milk. These findings support our findings among those aged 56-59. However, the authors did not stratify according to different age groups and they did not specify relevant substitutions for the changes in dairy intake. In a 9-week randomized intervention comparing commercially available low-fat yogurt with a soy pudding, reduced levels of some, not all, measured biomarkers of chronic inflammation and endotoxin exposure were observed in the low-fat yogurt group [34]. Overall, these findings support a benefit of yogurt products on risk of development of type 2 diabetes through mechanisms that may be mediated by changes in the gut microbiota, as suggested by others [35].

An advantage of specifying the food substitution is that it provides a clear interpretation of the results. We specified the association for increasing the intake of one dairy product subgroup at the expense of another. Our main model was Model 2, which also adjusted for other foods. However, it could be argued, for public health recommendations, that Model 1b, not adjusted for other foods, may be more relevant as this model did not constrain the underlying dietary pattern. For example, substitution of low-fat yogurt for skimmed milk may be associated with changes in the intake of other foods. We illustrated this using spider plots and generally found that those who increased their intake of low-fat yogurt also increased their intake of fruit, vegetables and coffee compared to those who did not change their intake of these dairy products. Indeed, after further adjustment for other foods (Model 2) some of the observed associations (Model 1b) disappeared. Although the magnitude of the RDs observed was rather small, around −0.9% to −0.5% for a substitution of 50 g/d for whole-fat yogurt with 50 g/d for milk products, this is not surprising, as we only modelled a single dietary change consisting of two different products.

We also investigated the modifying effect of the initial intake of the dairy product subgroup being replaced. Overall, the pattern of associations was similar among those with an initial high or low intake of the substituted dairy product. The low average initial intake of whole-fat dairy products in this cohort may partly explain the lack of a modifying effect. In our sex-stratified analyses we observed different patterns of associations between men and women. However, as there is little evidence of biological reasons for such differences, we interpret this with caution and encourage replication of this finding in other cohorts.

Future research should focus on randomized interventions and cohort studies with repeated measures of diet to evaluate these suggested differences between yogurt and milk products on risk of developing type 2 diabetes or risk markers hereof.

In conclusion, this study suggests a reduced risk of type 2 diabetes when replacing milk with whole-fat yogurt among men and women aged 56-59 years and an increased risk of type 2 diabetes when replacing skimmed milk for semi-skimmed milk among those aged 60-64 and 65-72, regardless of initial intake level.

## Data Availability

Data described in this article may be made available upon request pending on application to and approval by the Danish Cancer Society.

https://www.cancer.dk/dchdata/

## Abbreviations

BMI: Body mass index
CI: Confidence interval
FFQ: Food frequency questionnaire
RD: Risk difference

## Declarations

### Ethics approval

The study was approved by Regional Ethical Committees on human studies and by the Danish Data Protection Agency.

### Consent to participate

All participants gave written informed consent to participate in the study.

### Consent for publication

Not applicable.

### Code availability

Code is available upon request to the corresponding author.

### Authors’ contributions

The contributions were as follows-DBI, KO, ASDL, ETP and MUJ designed the study; DBI analysed the data supervised by ETP; DBI, KO, ASDL, JH, AT, TOK, ETP and MUJ interpreted the data and critically revised the manuscript; DBI wrote the paper; and DBI had final responsibility for the final content. All authors have read and approved the final manuscript.

## Acknowledgements

We thank the Danish Cancer Society, the staff at the Danish Diet, Cancer and Health study for collection of the data and the study participants for their contribution to the study. We thank data manager Nick Martinussen from the Danish Cancer Society for his large contribution with the harmonisation of the food frequency questionnaire data. We also thank data manager Katja Boll from the Danish Cancer Society for her work with the database and data administration.

